# Psycho-oncologists’ roles, tasks, and needs regarding requests for assisted suicide – a qualitative interview study on current experiences and future directions

**DOI:** 10.64898/2025.12.12.25342138

**Authors:** Zoe Henning, Isabelle Scholl, Pola Hahlweg

**Affiliations:** Department of Medical Psychology, University Medical Center Hamburg-Eppendorf, Hamburg, Germany

**Keywords:** assisted suicide, aid in dying, psycho-oncology, cancer, oncology, support

## Abstract

**Purpose:** Assisted suicide (AS) is a socio-political and healthcare challenge. In Germany, AS is legal, with further regulation pending. Previous German legislation drafts proposed involving psychosocial professionals like psycho-oncologists. These are experienced in supporting seriously ill patients and end-of-life decisions. However, there is a lack of studies on their role in AS. This study aimed to explore psycho-oncologists’ current and future roles, tasks and needs regarding AS requests.

**Methods:** We conducted a cross-sectional, qualitative interview study with psycho-oncologists in Germany. Inclusion criteria were current clinical activity as a psycho-oncologist and having talked to at least one patient about AS. Invitations to participate were distributed via email and social media using the study team’s network. Interested participants responded voluntarily (convenience sample). Data were analyzed using Practical Thematic Analysis.

**Results:** Twelve interviews were conducted (average length of 42 minutes). Participating psycho-oncologists (58% female, 50% up to 40 years, 100% psychology- or psychiatry-related professional background) primarily saw themselves as open-minded conversation partners, both currently and in the future. Opinions differed whether psycho-oncologists should assess capacity for free decision-making. The vast majority rejected participation in realizing AS. They opposed psycho-oncologists having to fulfill mandatory tasks in the context of AS. Key needs for engaging in AS-related work included clinical practice guidelines, legal clarity, and specific training opportunities.

**Conclusion:** This study provides initial insights into (potential) roles, tasks, and needs of psycho-oncologists in AS requests. It can serve as a basis for follow-up studies. Suitable structures and training opportunities should be developed.

## Background

Assisted suicide (AS) is an increasing, yet challenging phenomenon around the world[1, 2]. It refers to assisting a person in ending their own life, such as a physician prescribing a lethal medication that a patient self-administers[2]. The final act that leads to death has to be performed by the person dying[2]. Basing AS on a person’s right to autonomy and self-determination, the most important prerequisite for AS is the patient’s free decision[3, 4].

Supportive care aims to foster quality of life for cancer patients, also in end-of-life care[5]. It “involves a coordinated, person-centric, holistic (whole-person) approach, which should be guided by the individual’s preferences, and should include appropriate support of their family and friends”[5]. All of these characteristics apply to supporting requests for assisted suicide as well. Even though assisted suicide is ultimately about hastening one’s death, having the option of assisted suicide and being heard can simultaneously be about promoting quality of life.

When people seek AS, longitudinal and iterative processes unfold, including various participants, interactions, and decisions[6]. Thus, AS requests are a challenging situation in healthcare. Besides physicians, psychosocial healthcare professionals (HCPs) can be valuable in responding to AS requests, as AS processes include psychological and existential aspects[6-8]. Still, psychologists’ role in AS has been described as poorly established and underrepresented[9]. Potential AS-related roles and tasks for psychologists such as assessment of free decision-making capacity, communication, psychological assessment and support to patients and patient relatives, research and training, and public policy have been proposed[10, 11]. However, psychologists voiced limited confidence in assessing free decision-making capacity and a lack of specific training opportunities or guidelines for involvement in AS[9, 11, 12].

The current knowledge base remains limited. Research on AS rarely focused on psychologists and/or their roles, tasks, and needs in AS, but rather mentioned them in passing [e.g., [10, 13, 14]. Most studies were theoretical, literature-based, or quantitative without standardized questionnaires[10], omitting in-depth insights that can be deduced from empirical qualitative data. Furthermore, we assume that psycho-oncologists might be a sub-group of psychosocial HCPs especially relevant in the context of AS, as they have extensive experience in working with severely ill patients, end-of-life decision-making, and potentially AS[15]. To our knowledge, no scientific studies examined the role of psycho-oncologists in AS.

Thus, this study focused on psycho-oncologists’ current and potential future roles, tasks, and needs in AS requests, assessing them empirically and qualitatively from psycho-oncologists’ perspective.

## Methods

### Study design

We conducted a explorative cross-sectional qualitative interview study with psycho-oncologists [16]. We followed the Standards for Reporting Qualitative Research (SRQR)[17] (Supporting Information S1).

### Participants

We included psycho-oncologists with current clinical activity in Germany that had talked about AS with at least one patient. No additional exclusion criteria applied.

### Material

A semi-structured interview guide (Supporting Information S2) was developed based on the literature (cp. background section) and the research team’s expertise. It contained questions about actual roles and tasks related to AS requests (current experiences), roles and tasks they could take on (future directions), and what they need to be well equipped for these (needs). Participants also filled in a short demographic questionnaire (Supporting Information S3).

### Data collection

We used a convenience sampling approach, seeking maximum variation by study invitation and participant selection. Potential participants were invited across Germany between May and June 2024 via the Psycho-Oncology Working Group of the German Cancer Aid’s Network of Comprehensive Cancer Centers, the out-patient psychotherapy working group of the German Cancer Association’s Consortium for Psycho-Oncology, and a regional working group of psycho-oncologists. In addition, we sought participants via social media (Twitter/X, LinkedIn). Interested people received comprehensive information about the study from ZH by telephone, gave informed consent prior to participation, and were offered 25 Euros. Interviews were conducted by ZH via telephone or video conferencing (Zoom Video Communications, Version 6.1.11). Demographic questions were answered online via Limesurvey (LimeSurvey GmbH, *Limesurvey Community Edition*, Version 5.6.26).

Data saturation was iteratively discussed within the study team, using the concept of inductive thematic saturation[18]. Data collection stopped when data allowed enriching answers to the research questions within a feasible amount of time and resources.

### Data analysis

Interviews were audio recorded and transcribed using an offline Python script powered by a large language model that integrates multiple open-source libraries[19]. This script utilizes Whisper by OpenAI for speech recognition and pyannote.audio for speaker diarization[19]. Transcripts were thoroughly checked, revised and anonymized by ZH. Interview and demographic data were not linked.

Data was analyzed using Practical Thematic Analysis with slight adaptations[20]. It consisted of three iterative steps: reading, coding, and theming[20]. First, all transcripts were read to get familiar. Second, inductive codes were assigned as “labels for concepts that are directly relevant to the study objective”[20]. Initially, ten percent of the data were independently coded by two coders (ZH, PH, “consensus coding”). After consensus discussion, ZH continued coding and consulted PH in cases of uncertainty. Throughout the process, codes and memos were adapted and merged multiple times. Third, ZH identified meta-constructs (“themes”). In a joint session, ZH, PH, and IS discussed and revised the coding scheme until consensus was reached. Rater triangulation between ZH, PH, and IS was used to enhance analytic credibility. Demographic data was analyzed descriptively. We used MAXQDA software (VERBI GmbH, Version 22.2.0), Microsoft Excel (Microsoft, Version 2016), and SPSS (IBM, Version 29.0.1.0).

### Researchers’ characteristics

ZH is a female junior researcher and this study was her Master’s thesis. She had previously worked with PH on AS studies. PH and IS are female senior researchers and clinicians with comprehensive qualitative interview experience.

### Ethics approval and preregistration

This study was approved by the local ethics committee of the Center for Psychosocial Medicine at the University Medical Center Hamburg-Eppendorf (LPEK-0757) and pre-registered on Open Science Framework (https://doi.org/10.17605/OSF.IO/DSZ37).

## Results

In the following sections, we describe major findings (Figure 1). For the full coding scheme see Supporting Information 4.

**Figure 1.**
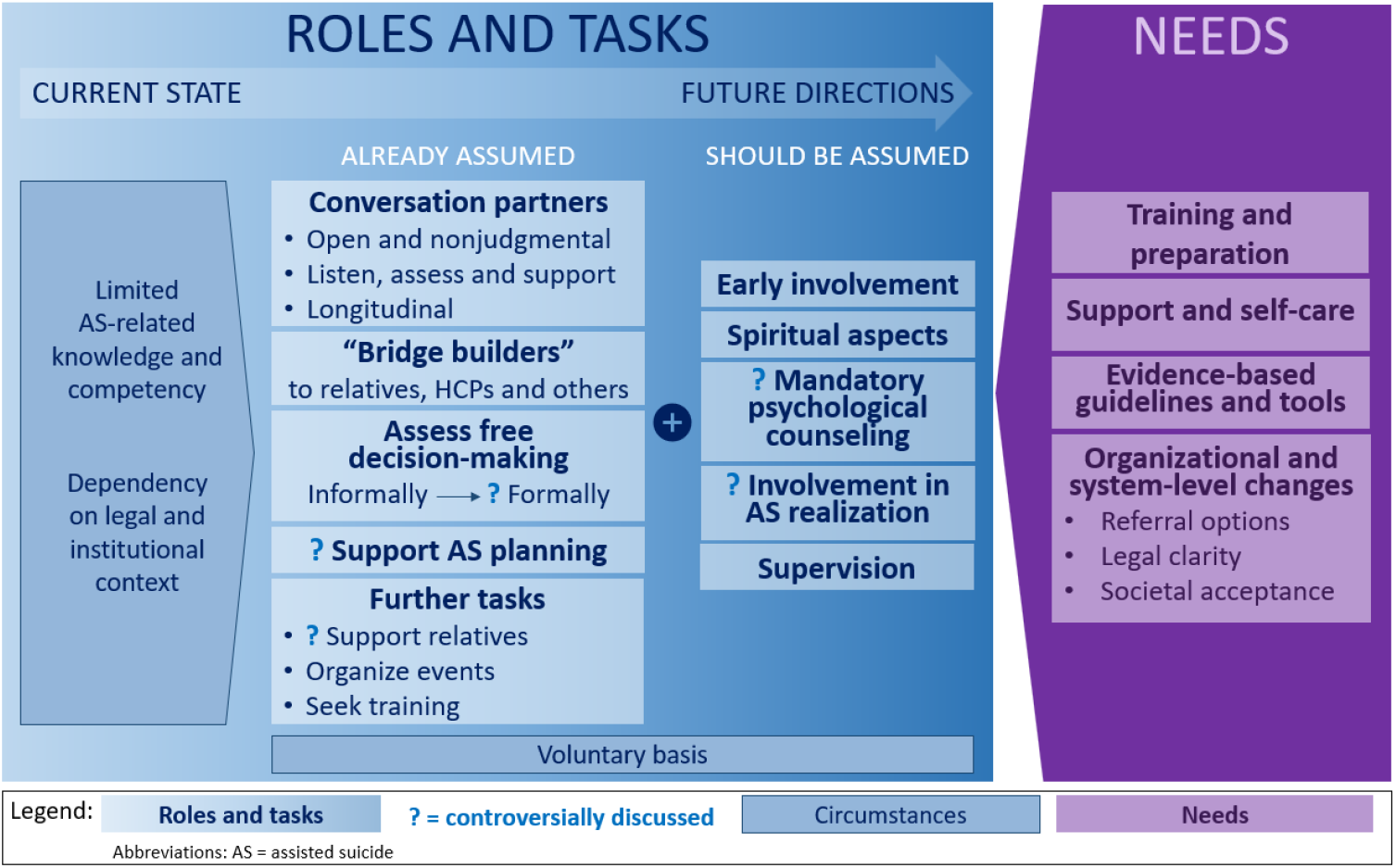
AS-related roles, tasks, and needs of psycho-oncologists.

### Sample characteristics

Twelve psycho-oncologists participated. Interviews lasted 42 minutes on average (SD=11; range 26-60). All participants had a professional background in psychology or psychiatry. Further demographic information is shown in Table 1.

**Table 1.**
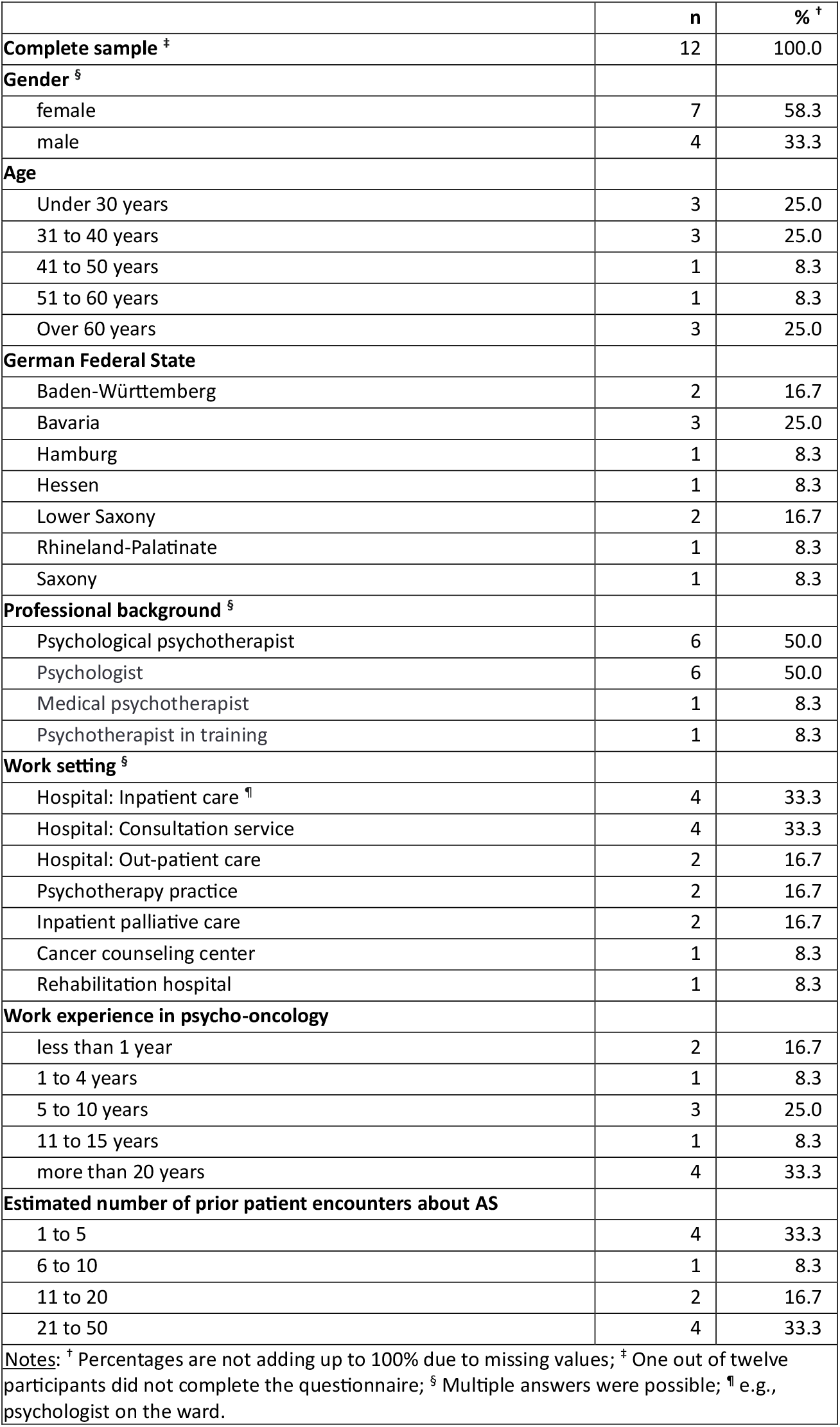
Sample characteristics.

|  | n | %<sup>†</sup> |
| --- | --- | --- |
| **Complete sample<sup>‡</sup>** | 12 | 100.0 |
| **Gender<sup>§</sup>** |  |  |
| female | 7 | 58.3 |
| male | 4 | 33.3 |
| **Age** |  |  |
| Under 30 years | 3 | 25.0 |
| 31 to 40 years | 3 | 25.0 |
| 41 to 50 years | 1 | 8.3 |
| 51 to 60 years | 1 | 8.3 |
| Over 60 years | 3 | 25.0 |
| **German Federal State** |  |  |
| Baden-Württemberg | 2 | 16.7 |
| Bavaria | 3 | 25.0 |
| Hamburg | 1 | 8.3 |
| Hessen | 1 | 8.3 |
| Lower Saxony | 2 | 16.7 |
| Rhineland-Palatinate | 1 | 8.3 |
| Saxony | 1 | 8.3 |
| **Professional background<sup>§</sup>** |  |  |
| Psychological psychotherapist | 6 | 50.0 |
| Psychologist | 6 | 50.0 |
| Medical psychotherapist | 1 | 8.3 |
| Psychotherapist in training | 1 | 8.3 |
| **Work setting<sup>§</sup>** |  |  |
| Hospital: Inpatient care<sup>¶</sup> | 4 | 33.3 |
| Hospital: Consultation service | 4 | 33.3 |
| Hospital: Out-patient care | 2 | 16.7 |
| Psychotherapy practice | 2 | 16.7 |
| Inpatient palliative care | 2 | 16.7 |
| Cancer counseling center | 1 | 8.3 |
| Rehabilitation hospital | 1 | 8.3 |
| **Work experience in psycho-oncology** |  |  |
| less than 1 year | 2 | 16.7 |
| 1 to 4 years | 1 | 8.3 |
| 5 to 10 years | 3 | 25.0 |
| 11 to 15 years | 1 | 8.3 |
| more than 20 years | 4 | 33.3 |
| **Estimated number of prior patient encounters about AS** |  |  |
| 1 to 5 | 4 | 33.3 |
| 6 to 10 | 1 | 8.3 |
| 11 to 20 | 2 | 16.7 |
| 21 to 50 | 4 | 33.3 |
| **Notes:** <sup>†</sup> Percentages are not adding up to 100% due to missing values; <sup>‡</sup> One out of twelve participants did not complete the questionnaire; <sup>§</sup> Multiple answers were possible; <sup>¶</sup> e.g., psychologist on the ward. |  |  |

### Current AS-related experiences of psycho-oncologists

#### Roles

Participants primarily viewed themselves as *open, non-judgmental conversation partners*, providing safe spaces to discuss any topic, including AS.

> *“I see my role as neutrally examining the situation together with the person, without making any judgment, and observing their actual stance on the matter. So, a very inquisitive role, with no intention of steering them in any direction*.*”* (T03)

Some described themselves as *longitudinal supporters* in AS decision-making, assisting patients over multiple discussions through possible ambivalence and potential decision changes. Another AS-related role was being *“bridge builders”* by *involving relatives* and *collaborating with HCPs and others*. Participants described their current role as *talking with patients about AS*, but *not being present during the realization* of AS.

#### Conversational tasks

Participants reported *various conversational tasks* in response to AS requests. AS discussions were mostly *patient-initiated* and *heard and taken up* by psycho-oncologists. Few participants had *proactively raised* the topic of AS with patients. When talking about AS, psycho-oncologists *explored AS wishes, suicidality* and *free decision-making*. While suicidality assessment was described as formal and could trigger mandatory consequences, the exploration of free decision-making was informal.

> “*When exploring, it’s important to me that it’s not suicidality. Especially when it is a treatable aspect of depression, as it would then not be correct. Because it would not be self-determined, but illness-determined*.*”* (T05)

Furthermore, participants described to *provide neutral information about AS, discuss alternatives to A*S, *refer patients to additional support structures*, and *support during AS planning*. One participant emphasized the importance of *balancing* respect for *patient autonomy* and *protecting life* in dialogue. Psycho-oncologists also *offer psychotherapeutic interventions* and *stay in contact, if a patient is already registered for AS*.

#### Further tasks

Psycho-oncologists also *support patient relatives* during and/or after AS.

> *“ So that it doesn’t represent an unreasonable burden within the relationship for his wife, but rather something she can live with. That he wants this [AS], and that she can have the feeling, ‘I support you, but at the same time, I want you to live as long as possible*.*’”* (T06)

Some had *organized AS-related professional events*, such as university courses or professional training sessions. Participants also described being proactive in *seeking out education and training* for themselves.

#### (Limited) knowledge and competencies

Many participants felt *inadequately informed* about key aspects of AS, such as terminology and legal requirements, resulting in a sense of unpreparedness for AS counseling or free decision-making assessment. Insecurity was particularly pronounced regarding AS requests from *vulnerable groups*, such as individuals with intellectual disabilities or children. In contrast, many respondents felt *well-prepared for other aspects of AS-related work*, such as supporting decision-making processes.

#### Legal and institutional context

Participants highlighted their ability to undertake AS tasks being highly *setting- and context-dependent*. For example, short patient stays in hospitals often precluded psycho-oncologists from adequately supporting AS decision-making. S*uitable spaces* for AS discussions might not be available, especially in acute care settings, and a *lack of financial reimbursement* for some AS-related tasks was mentioned for some settings. Furthermore, many participants criticized the *lack of clear guidelines and policies* on the institutional and system level.

> *“Right now, it’s kind of like the Wild West regarding what is or isn’t possible with AS. […] And here at [hospital], we barely have any guidelines. There’s no SOP [Standard Operating Procedure], nothing in the quality management manual*.*”* (T02)

In some institutions, AS-related conversations with patients were *explicitly prohibited* (*“Even talking about it at the bedside is grounds for a warning in our institution. “*, T08).

The extent of *collegial exchange* on AS-related experiences also varied. Several participants reported *limited or no discussions with colleagues*, others had had *meaningful exchanges*.

### Future directions for AS-related tasks and roles of psycho-oncologists

Participants reported *uncontroversial* (i.e., agreed among participants) and *controversial* (i.e., differing opinions) future roles and tasks. Furthermore, they mentioned roles and tasks psycho-oncologists *already undertake* and should keep and those they *should add* in the future.

In general, participants emphasized psycho-oncologists’ involvement in AS-related tasks having to remain *voluntary* and *free from legal or employer-imposed mandates*. Psycho-oncologists should be able to decide this case-by-case.

#### Uncontroversial roles and tasks

Many of the current roles and tasks were reiterated. These involved being open conversation partners and “bridge builders”, exploring AS wishes, providing neutral information about AS, discussing alternatives to AS, making referrals, and offering psychotherapeutic interventions. Uncontroversial tasks to add in the future included *being involved early* in AS considerations, *addressing spiritual and religious concerns*, and *offering supervision* on AS to other HCPs.

#### Controversial roles and tasks

Three current roles and tasks were controversially discussed for the future: *assessing free decision-making, supporting during AS planning*, and *supporting patient relatives*. Many viewed assessing free decision-making as a task psycho-oncologists should undertake. Others opposed the idea of assuming evaluative or certifying roles, suggesting these tasks are better suited to psychiatrists or AS organizations.

> *“If I were to be the person who ultimately evaluates free decision-making and gives the final approval […] I think that would be a reason for me to quit. […] Some individuals may want to specialize in this area, but if I were forced to do it, it would cross a line for me*.*”* (T11)

Support in AS planning was primarily deemed physicians’ responsibilites, with some aspects possibly being psycho-oncological tasks as well. In contrast, most promoted supporting relatives, while one participant opposed this role.

Furthermore, opinions were divided on the necessity of *mandatory psychological counseling* regarding AS. *AS realization* was considered primarily the responsibility of physicians and psycho-oncologists’ involvement was questioned by many (*“I think my role would always be that of a conversation partner […], but not for the actual realization*.*”*, T12). However, two participants could imagine involvement during AS realization in exceptional cases.

### Psycho-oncologists’ needs regarding AS-related work

#### Training and preparation

Participants emphasized the need for *specific training* to address AS requests and reduce professional uncertainty. This training could be organized as standalone AS-focused courses or be integrated into existing programs. Practical *workshops on AS communication skills* were asked for. In addition, participants stressed the importance of psycho-oncologists *reflecting on their personal attitudes* toward AS. This is necessary to avoid personal biases influencing therapeutic interactions, as AS touches on deeply held personal and ethical convictions.

#### Support and self-care

Psycho-oncologists highlighted that they need support systems themselves, including *AS-specific supervision and collegial exchange*, to manage the emotional burden. Furthermore, participants need *self-care* as part of their work. A team culture that accepts and acknowledges self-care and sufficiently flexible schedules would be necessary. Self-care was important to participants for their own mental health.

#### Evidence-based guidelines and tools

*Evidence-based guidelines and tools* should include protocols for assessing free decision-making, providing AS information, and documenting AS discussions. Practical resources such as patient brochures would also be helpful.

> „*I would also like to have a guideline […] that outlines: What alternatives to assisted suicide can I present in psychoeducational conversations? What [AS] associations exist? How do they typically work? What are the legal foundations? A brochure like that would provide good orientation and could be something to share with patients. And a guideline specifying which key areas should definitely be addressed in conversations*.*” (T09)*

To develop such resources and ensure they are evidence-based, participants underscored the need for further AS-related research.

#### Institutional and healthcare system changes

Some participants reported a need for institutional adjustments, including *freedom from restrictive institutional policies*. Participants also reported the need for a *well-organized team with good communication. Confidential spaces* and *adequate time* for patient interactions were described as necessary (“*I would think that enough time and in-depth conversation spaces are needed […]*.”, T11).

In addition, *financial barriers*, such as limited reimbursement for AS-related tasks, need to be addressed. Importantly, psycho-oncologists should receive compensation independently of the realization of AS to maintain professional neutrality.

Establishing *clear referral pathways* regarding AS requests was considered essential, within and between institutions („*We can refer to structures, but these structures are actually not there yet. And that makes it problematic*.*”*, T05). A need for *contact persons responsible for AS* who open up discussion spaces was voiced. Participants furthermore asked for AS-related *information sessions for patients*. Such contacts would clarify responsibilities and provide structured support for both psycho-oncologists and patients.

Participants need current AS-related legal uncertainties to be addressed. *Clear legal requirements* could protect professionals from potential liabilities.

> *“I believe really concrete guidelines and recommendations would be important […]. That it is all very specific, so that one doesn’t have to be afraid of facing difficulties when doing this or helping with it [AS]*.*”* (T10)

The importance of greater *societal acceptance of AS* and AS-related work was highlighted. Shifting AS discussions out of the “*forbidden zone*” (T01) could foster broader understanding and empathy. This shift could positively impact patients and ease the burden on psycho-oncologists.

## Discussion

This qualitative study explored psycho-oncologists’ current and potential future roles, tasks, and needs regarding AS-requests in Germany. Currently, psycho-oncologists predominantly described themselves as open, non-judgmental and dependable conversation partners, offering a safe space for exploration, information, clarification, balancing and possibly intervening. Supporting patient relatives, offering AS education and training opportunities, and educating oneself were mentioned as further tasks. However, participants reported limited current knowledge and competencies regarding AS and being influenced by institutional and legal contexts. They emphasized that AS involvement must remain voluntary. In the future, they still saw themselves in a mostly as supportive conversation partners, with few wanting an evaluative role. Early involved in AS processes, considering spiritual aspects, and offering supervision to HCPs were uncontroversial additional tasks. If psycho-oncologists should formally assess free decision-making capacity, be involved in AS planning and possibly realization, and if psychological AS counselling should be mandatory, was controversially discussed. To be well-equipped for AS-related work, participants would need specialized training opportunities, supervision and team support, self-reflection and self-care, practice guidelines, and organizational and system changes such as referral options, legal clarity, and societal acceptance of AS.

Our results suggest that, in the context of AS, psycho-oncologists predominantly see themselves as dependable conversation partners offering a safe space, both now and in the future. In our study and psycho-oncology in general, corresponding core tasks include in-depth exploration, decision-making support, mediating between patients, their relatives and other HCPs, and psychological interventions[21].

Regarding evaluative AS tasks, most psycho-oncologists in our sample were hesitant. The fact that these assessments have implications on life and death make the stakes very high. Hesitancy might be fueled by uncertainty regarding assessment procedures and requirements, lack of training, or institutional restrictions. This is especially noteworthy, as some participants had sought specific education and training and organized AS-related events, suggesting our sample might have been better informed than the general population of psycho-oncologists. Psycho-oncologists and psychologists are often depicted as supporters, not evaluators. As certain psychology subfields, e.g. transplantation[22] or forensic psychology[23], are commonly undertaking evaluative roles, comparing their processes to AS could be an interesting next step.

How to assess free decision-making has been widely discussed as a challenge, far beyond the field of psycho-oncology[24]. If AS processes are to be offered with high quality, sound assessment of free decision-making, the most important prerequisite for AS, needs to be established. Scientific development of measurement tools and assessment interviews are crucial. Psycho-oncologists and psychologists bring clinical experience and research competencies (e.g., psychometrics) to the table.

Participants of this study emphasized that any involvement of psycho-oncologists in AS processes has to be on a voluntary basis and cannot be mandated. This aligns with the general argument for a right to conscientious objection regarding AS involvement[25].

Limited confidence regarding certain tasks and lack of knowledge, competence, or training repeatedly emerged from our data. Thus, specialized training and the development of standards was named as essential to offer good care for the challenging topic of AS. Psycho-oncologists in our sample saw themselves both on giving and receiving such training and supervision. They also called for organizational and system changes to alleviate legal, institutional, and procedural uncertainties.

Participants in this study wanted high-quality and accessible AS processes for their patients, but not necessarily offer all of it themselves. They called for effective referral pathways. They also emphasized the need for good collegial exchange and team support, since AS involvement should not be a one-person task[6].

Our results for psycho-oncologists mostly align with other studies regarding AS-related roles and tasks of psychologists (cp. background section)[8, 11]. Nevertheless, our study highlighted some additional aspects: First, it emphasized the necessity for AS involvement to be voluntary. Second, psycho-oncologists called for early and longitudinal involvement in AS processes. Third, spiritual considerations were mentioned as an AS-related task for psycho-oncologists. This aligns with tasks described for psychologists in palliative care[14, 26]. Fourth, influences on the organizational and system-level were described. Fifth, albeit controversially, psycho-oncologists involvement in AS planning and realization was mentioned.

### Strengths and limitations

To our knowledge, this is the first study exploring psycho-oncologists involvement in AS processes. The structured inductive analysis and rater triangulation are major strengths of this study. Convenience sampling might have led to participants being more open to AS than the general population of psycho-oncologists. However, generalizability is not the primary aim of qualitative research[16]. Nevertheless, confirmation of the results is needed. Furthermore, our interview guide being informed by prior research might have made alignment between our results and prior research more likely. Nevertheless, we asked open-ended questions and found additional new aspects in our data.

### Practice implications

As this was an exploratory study, practice implications have to be drawn with caution. Confirmatory research is needed prior to firm recommendations. We suggest subsequent quantitative studies with robust sampling, including psycho-oncologists who are ambivalent or opposing AS, and exploring other perspectives (e.g., patients who discussed AS with psycho-oncologists).

Nevertheless, common psycho-oncological roles and tasks, such as being open and supportive conversation partners, have been found to be applicable to the context of AS.Nevertheless, specialized AS training was called for. The crucial task of assessing free decision-making was associated with uncertainty and limited subjective competence. Thus, developing reliable standards, assessment procedures, and appropriate training would be beneficial. As the concrete planning and realization of AS differs from common psycho-oncological tasks, further clarification regarding psycho-oncologists potential involvement is needed. Finally, organizational and system changes (e.g., clarifying legal regulations, clinical practice guidelines, institutional support) need to be addressed on higher levels.

## Conclusion

This study provided initial insights into roles, responsibilities, and needs of psycho-oncologists in AS requests. Psycho-oncologists already undertake tasks for AS requests and some would be willing to take on further tasks, if their needs are addressed. While they currently predominantly described themselves as supportive conversation partners, we found potential for additional roles and tasks such as formally assessing free decision-making or supporting the planning and realization of AS. Voluntariness as the basis for undertaking any AS-related task was crucial. Meeting psycho-oncologists needs would require, among other things, clarifying legal and institutional regulations, establishing practice guidelines, and expanding training opportunities. Furthermore, suitable structures that allow AS referrals to specialists should be established. The results of this study can inform subsequent research and health policy developments.

## Supporting information

SuppInfo1_SRQR-Checklist

SuppInfo2_Interview Guide

SuppInfo3_Demographic Questionnaire

SuppInfo4_Coding Scheme

## Declarations

## Acknowledgements

We greatly appreciate all psycho-oncologists who participated in this study. We furthermore thank Max Grohmann for programming and setting up the transcription software. Last but not least, we thank Prof. Dr. Friedemann Geiger who co-supervised ZH’s Master’s thesis.

## Author contributions

ZH contributed to conceptualization, data curation, formal analysis, investigation, methodology, project administration, visualization, and writing the original draft. IS contributed to conceptualization, formal analysis, supervision, and writing by reviewing and editing. PH, the principal investigator of this study, contributed to conceptualization, formal analysis, methodology, project administration, supervision, visualization, and writing the original draft.

## Data availability

The qualitative data collected and analyzed during the translation process (in German) are available from the corresponding author on reasonable request. Signing a data use/sharing agreement will be necessary, and data security regulations both in Germany and in the country of the investigator, who proposes to use the data, must be complied with. Preparing data sets for use by other investigators requires substantial work and is thus linked to available or provided resources.

## Use of generative AI and AI-assisted technologies

During the preparation of this work, the authors used DeepL in order to improve language and readability. As described in the methods section, they used an offline Python script powered by a large language model that integrates multiple open-source libraries to initially transcribe the audio-recorded interviews. After using these tools, the authors carefully reviewed and edited the content and take full responsibility for the content of the transcripts and the publication.

