## Supplementary material for "Psycho-oncologists’ roles, tasks, and needs regarding requests for assisted suicide – a qualitative interview study on current experiences and future directions": SuppInfo2_Interview Guide

### Supporting Information 2 | Semi-structured interview guide

#### Preparations

- Schedule an appointment and clarify the setting (phone/video call)
- Has the consent form been completed?
- Are the recording devices ready and charged?

#### Interview Phase 0: Introduction

*ZH calls the participant at the agreed time.*

- Hello, my name is Zoe Henning from the University Medical Center Hamburg-Eppendorf. Am I speaking to [name of participant]?
- I am calling for our scheduled telephone interview. Thank you again for agreeing to participate. The interview will last approximately **45–60 minutes**. I have already received your **consent form**. Do you have any **questions** before we start? […] All right, then we will now begin the interview. As agreed, I will start the audio recording. Okay?

➔ **TURN ON AUDIO RECORDING DEVICE**

- To start, I would like to ask you once again, while the recording is running, whether you consent to participate in the study. […]
- As you know, in this interview, we will be discussing the role of psycho-oncologists in the context of assisted suicide (AS). The aim of the study is to examine the (potential) roles and responsibilities of psycho-oncologists in AS requests. Through these interviews, we aim to understand what tasks you currently undertake in AS cases, what tasks you could take on in the future, and what you would need for such involvement. We are specifically interested in the role of psycho-oncologists in AS, not in any potential medical activities related to the process.
- I will begin by providing a brief introduction and asking you some general questions about your professional background. After that, we will proceed with the questions on AS.
- During the interview, I will ask you **questions step by step**. Please feel free to share anything that comes to mind. It is important for this interview that you feel comfortable **expressing your opinions and experiences** openly and honestly. There are **no right or wrong** answers. You are welcome to share both **critical and positive** perspectives.
- Your data and statements will be treated **confidentially** and analyzed **anonymously**. As you know, the recording will be **transcribed** after the interview. During this process, all names and identifying characteristics will be removed so that **nothing can be traced back to you as an individual**. However, I want to emphasize that it is entirely up to you, which

information you choose to share. You can stop the interview at any time, if you feel uncomfortable.

##### Introduction:

Before we begin, I would like to clarify some key terms, even though you are likely already familiar with them.

- In this interview, we will be discussing AS, which refers to a person choosing to end their own life voluntarily with assistance from another person. For example, a physician could provide access to medication that allows the person to end their own life. The goal of AS is the person's death. AS is legal in Germany.
- This interview does not concern euthanasia, which refers to a situation where the assisting person actively performs the final act, such as administering a lethal injection. Euthanasia is illegal in Germany.
- Additionally, this interview does not cover symptom relief or treatment limitation at the end of life, as these medical measures do not have the direct goal of causing death.
- Today, our focus is on AS. Is this distinction clear? […]

##### Introductory Questions on Professional Background:

First, I would like to learn more about your professional role.

- Please briefly describe your work context, in which you have encountered or might encounter AS.

#### Interview Phase 1: Current Experiences

In the first part, I would like to talk to you about your past experiences.

- Have you ever discussed AS with patients in your professional context or even been involved in supporting a patient through AS?
    - If yes: With approximately how many individuals have you discussed AS? How many patients have you accompanied during AS?
- What role or responsibilities have you taken on as a psycho-oncologist with these patients? *[If not mentioned, ask more specifically:]*
    - What exactly did you do in [XX] cases?
    - What did you discuss with your patients regarding AS?
- Have you observed colleagues taking on other tasks in AS cases?
- What challenges or uncertainties do you currently face in relation to your role in AS?

#### Interview Phase 2: Future Directions

Now, I would like to discuss what role you believe psycho-oncologists should or could have in AS.

- How beneficial do you find the involvement of psycho-oncologists in AS?
- What tasks should or could psycho-oncologists take on in AS in the future? What would the ideal situation look like?

  *[If not mentioned, ask further:]*
  - To what extent should or could psycho-oncologists be involved in **AS counseling**?
  - To what extent should or could psycho-oncologists be involved in **evaluating AS requests**?

    *[e.g., determining self-responsibility and differentiating from suicidality due to mental illness]*
  - To what extent should or could psycho-oncologists be involved in **planning and carrying out AS**?
  - To what extent should or could psycho-oncologists be involved in **aftercare** following an AS?
  - **For whom** should or could psycho-oncologists offer **psychological support** in the context of AS?

#### Interview Phase 3: Needs

Finally, I would like to discuss what you or your psycho-oncology colleagues would need in order to effectively take on the tasks you mentioned regarding AS.

- How well prepared do you currently feel to take on these tasks?
- What conditions would need to be met, or what would you require to carry out these tasks effectively?
  - What would be personally helpful or supportive for you?
  - … and what would be personally challenging or difficult for you?

#### Interview Phase 4: Conclusion

**Summary:**

- Looking back at everything we discussed today, which aspects are most important to you?
- Is there anything we haven't covered that you would like to add?

#### Closing:

- We have now reached the end of the interview. I sincerely thank you for this inspiring and insightful conversation. I will now send you a link via email to a short anonymous online questionnaire. I kindly ask you to complete it so that we can describe the group of participants. At the end of the questionnaire, you will find a link to claim the financial compensation for your participation.
- If you have any questions after the interview, please feel free to contact us via email. I wish you a great day. Goodbye!

*Turn off audio recording device*
