## Supplementary material for "Psycho-oncologists’ roles, tasks, and needs regarding requests for assisted suicide – a qualitative interview study on current experiences and future directions": SuppInfo3_Demographic Questionnaire

### Supporting Information 3 | Demographic Online Questionnaire for Interview Participants (English translation)

Note: This is an English translation. The original questionnaire was in German.

**Please check the appropriate box(es):**

1. What gender do you identify as? *(Multiple answers possible)*
   - ☐ Female
   - ☐ Male
   - ☐ Non-binary/diverse
   - ☐ Other or prefer not to say

2. Which age group do you belong to?
   - ☐ Under 30 years
   - ☐ 31 to 40 years
   - ☐ 41 to 50 years
   - ☐ 51 to 60 years
   - ☐ Over 60 years
   - ☐ Prefer not to say

3. In which federal state do you work?
   - ☐ Baden-Württemberg
   - ☐ Bavaria
   - ☐ Berlin
   - ☐ Brandenburg
   - ☐ Bremen
   - ☐ Hamburg
   - ☐ Hessen
   - ☐ Mecklenburg-Western Pomerania
   - ☐ Lower Saxony
   - ☐ North Rhine-Westphalia
   - ☐ Rhineland-Palatinate
   - ☐ Saarland
   - ☐ Saxony
   - ☐ Saxony -Anhalt
   - ☐ Schleswig-Holstein
   - ☐ Thuringia
   - ☐ Prefer not to say

4. In which setting do you currently work as a psycho-oncologist? *(Multiple answers possible)*
   - ☐ Hospital: Outpatient care
   - ☐ Hospital: Inpatient care (e.g., ward psychologist, liaison service)
   - ☐ Hospital: Consultation service
   - ☐ Cancer counseling center
   - ☐ Psychotherapy practice
   - ☐ Inpatient palliative care
   - ☐ Outpatient palliative care
   - ☐ Rehabilitation hospital
   - ☐ Prefer not to say

5. What is your professional background?
*(Multiple answers possible)*

- ☐ Clinical psychologist*
- ☐ Psychologist
- ☐ Physician trained in psychotherapy**
- ☐ Physician
- ☐ Educator
- ☐ Prefer not to say
- ☐ Other professional background (please specify in the comment field): ____________________

6. How many years of work experience do you have in psycho-oncology?

- ☐ Less than 1 year
- ☐ 1 to less than 5 years
- ☐ 5–10 years
- ☐ 11–15 years
- ☐ 16-20 years
- ☐ More than 20 years
- ☐ Prefer not to say

7. With how many of your psycho-oncological patients did you discuss assisted suicide? *(An approximate estimate is sufficient.)*

- ☐ 0
- ☐ 1-5
- ☐ 6-10
- ☐ 11-20
- ☐ 21-50
- ☐ More than 50
- ☐ Prefer not to say

<u>Notes:</u>
* In German: ‚psychologische:r Psychotherapeut:in', i.e., a licensed clinical psychologist
** In German: „ärztliche:r Psychotherapeut:in", i.e., a physician with additional training and a license to provide psychotherapy
