## Supplementary material for "Psycho-oncologists’ roles, tasks, and needs regarding requests for assisted suicide – a qualitative interview study on current experiences and future directions": SuppInfo4_Coding Scheme

### Supporting Information 4 | Coding scheme including codes, code descriptions, quotes, overall code frequencies, and frequencies of documents per code

<u>Notes:</u> These results are based on 12 interviews and should be interpreted as purely exploratory. Psych = psycho-oncologist(s), pat = patient(s), n (doc) = number of documents with this code, n (code) = number of codes across all documents, (TXX, XX) = number of the transcript and line number within the transcript.

| ID | Code | Description | Quote | n (doc) | n (code) |
|---|---|---|---|---|---|
| **Research Question (RQ) 1: What roles and tasks do psycho-oncologists currently undertake in requests for assisted suicide? (current state)** | | | | | |
| **A1** | **Psych take on different roles in response to AS requests** | | | | |
| A1.1 | Psych are open conversation partners | Psych are open conversation partners for pat on AS, responding nonjudgmentally when the topic arises and creating spaces for self-reflection on ambivalences and decision-making. | I see my role as neutrally examining the situation together with the person, without making any judgment, and observing their actual stance on the matter. So, a very inquisitive role, with no intention of steering them in any direction or not steering them – just ensuring that the process works. (T03, 32) | 10 | 24 |
| A1.2 | Psych are longitudinal supporters | Psych support AS decision-making processes longitudinally, remaining open to ambivalence and changes in decisions throughout multiple conversations. | I frequently experience that it is a long process. I currently have a patient who is becoming more advanced in his decision, and it might be possible in the next few days or weeks for him to make a definitive decision. I've been talking to him about it [assisted suicide] for almost a year now. From my perspective, I find it incredibly valuable that there is space in the process for the various ambivalences that arise. (T12, 37) | 4 | 7 |
| A1.3 | Psych are "bridge builders" | | | | |
| *A1.3.1* | *Psych involve relatives* | Psych involve patients' relatives in the AS decision-making process, either mentally or personally. Mentally, they discuss relatives' views on AS, its potential impact, and how to manage conflicting needs. Personally, involving relatives helps them understand the decision and become part of the decision-making process. | And it was more my task to 'get my will across in a partnership-friendly way.' So that within the relationship, it wouldn't impose an unreasonable burden on his wife, but that she could live with the fact that he wants this, and that she could feel, 'I support you, but at the same time, I want you to live as long as possible.' (T06, 24) | 6 | 15 |

| | | | | | |
|---|---|---|---|---|---|
| *A1.3.2* | *Psych collaborate with HCPs and others* | Psych collaborate with others, either being consulted by them (e.g., nursing staff, physicians, self-help associations) or involving them for support (e.g., hospice, palliative care, psychiatrists). | And in the other case, it was not just me looking at the patient alone, but the palliative care team and the psychiatrist also came. (T09, 28) | 6 | 12 |
| A1.4 | Psych talk about AS, but are not realizing AS | Psych discuss AS with pat but have not been involved in its execution. Reasons include viewing it as not part of their role, the setting not allowing for such support, or not reaching that point with the pat. | I save myself from uncertainties by clearly defining my role as a conversation partner who does not actively support or realize this process. (T12, 41) | 8 | 17 |
| **A2** | **Psych take on various conversational tasks in response to AS requests** | | | | |
| A2.1 | AS conversations are mostly initiated by pat, rarely by psychologists | | | | |
| *A2.1.1* | *Psych hear the topic, when pat bring up AS* | The conversation about AS is initiated by the patient, often indirectly through hints. Psychologists engage with the topic but do not address it proactively. | Actually, the topic is always brought up by the patients. I've never raised it myself. I don't think it would be appropriate, not appropriate at all. And it's often the case that the patients say, 'Well, I don't even know if I want to do this under these conditions, if I even want to live anymore.'" (T03, 36) | 8 | 12 |
| *A2.1.2* | *Psych proactively raised the topic of AS with pat* | When the topic of AS is on the table, some psychologists bring up the option of AS on their own. | For example, it helped me to bring up this topic as well. I asked the person what they thought of this option, how they felt about it, and whether they could imagine having an assisted suicide. It quickly became clear that the person did not want that at the moment. (T01, 32) | 3 | 3 |
| A2.2 | Psych explore AS wishes | Psych explore and reflect with patients on their AS wishes (e.g., considering reasons and influencing factors such as current stressors, symptom burden, impact on family, fears, conditions of origin, motivation, needs, pros and cons, completeness of considerations, resources, and social environment) to support patients in self-reflection and decision-making | And I see my role as asking questions about how the thoughts on this look exactly, how someone thinks about it, in order to explore it in a classical psychological way. [...] It's often about ambivalence, and I think then I typically give space to the ambivalence, to all the options that come into question, to the impulses, the needs, and try to deepen that, to explore what needs lie behind it. So, a lot of exploring and supporting someone in their self-reflection. (T12, 33) | 10 | 28 |

| | | | | | |
|---|---|---|---|---|---|
| A2.3 | Psych explore, document, and report suicidality | Psych explore suicidal thoughts and inquire about suicide plans. They report expressed suicidal thoughts and plans to the team, document them, and initiate further steps for suicide prevention, if necessary. | And if someone has a suicide plan, if it goes beyond just the wish, it is my responsibility to pass that on. I always consult with the medical staff in any case. We need to know if someone has suicidal wishes or thoughts, and how intense they are. Can this patient refrain from acting on them in the current situation in the hospital? Does this need to be included in the medical report? It must be documented very carefully (T08, 28) | 7 | 11 |
| A2.4 | Psych informally explore free decision-making | Psych explore whether the patient's capacity for free decision-making is impaired, particularly due to mental disorders and associated suicidality. | It's important to me when I explore that it's not a case of suicidality. That is especially the case when this [AS wish] appears as a treatable aspect of depression, as it would then not be correct. Because it would not be self-determined, but illness-determined and that is not my view of [assisted] suicide, that there are no alternatives. It must be a free choice." (T05, 20) | 4 | 5 |
| A2.5 | Psych provide neutral information about AS | Psych provide neutral and unbiased information about the conditions and possible process of AS. | There are patients who don't know how this works, so I try to explain as neutrally as possible how it goes. Or I talk about other patients who have done it this way or that. (T06, 32) | 6 | 7 |
| A2.6 | Psych discuss alternatives to AS | Psych discuss alternatives to AS with pat, including options like palliative care, psychiatric care facilities, or intensive psychological support—some of which may aim to improve pat' quality of life or strengthen their will to live. | When I talked to patients about it, they said the topic was off the table. They were rather grateful to have discussed palliative care options again – that there is, for example, the possibility of palliative sedation at the end of life. It was more a lack of knowledge among patients who were in distress, wondering what to do at the end of life when the suffering becomes unbearable. (T09, 32) | 8 | 17 |
| A2.7 | Psych refer pat to additional support structures | Depending on patients' needs, psychologists refer them to appropriate services (e.g., right-to-die organizations, support groups, psychotherapy, psychiatry, palliative care, physicians, or care facilities), and may also guide them in finding information about AS, such as by recommending online research. | Contact with patients is initiated when they wish for it – when they say, 'This is my path, I have made my decision, this is what I want.' Then, such a contact can be passed on. Before that, patients might be referred to organizations – there's Dignitas, for example – organizations that engage with the topic and can offer a good approach to assisted suicide. (T01, 40) | 10 | 17 |
| A2.8 | Psych support pat during AS planning | Psych are involved in the conceptual planning of AS by discussing with pat, for example, appropriate timing and potential providers of assistance. | And when I provided information in that regard – like when someone said they were planning to go to Switzerland, which happened quite often – I would ask, | 4 | 6 |

| | | | | | |
|---|---|---|---|---|---|
| | | | 'When would be the right time to go to Switzerland? (T09, 32) | | |
| A2.9 | Psych focus on balancing respect for pat autonomy and protecting life | In their professional stance, as well as in navigating societal and legal positions on AS, psych face the challenging task of balancing the protection of life with respect for autonomous end-of-life decisions. | I would highlight both sides: On the one hand, there is a legitimate interest among many patients in having the option to end their lives responsibly and without resorting to undignified means, like taking a rope or something similar. That is truly undignified. They deserve much understanding, and we should create opportunities to help them. On the other hand, I have concerns that this could be interpreted too freely, and that we are working with a concept of autonomy that is ideologically driven, one that emphasizes our ability to be autonomous too much. There is also the risk that social pressure could emerge, such as: 'This option exists, so why not use it?' This could be supported by compassion, but also by financial interests or ulterior motives. That is what I fear. (T06, 80) | 1 | 8 |
| A2.10 | Psych offer psychotherapeutic interventions | Psych apply psychotherapeutic interventions when needed, such as stabilizing, resource-oriented, and supportive work with pat. They also use appropriate interventions for psychological symptoms like anxiety or depression. | My role is primarily exploration and support with severe depressive thoughts, and psychotherapeutic intervention for significant anxiety. (T08, 28) | 6 | 9 |
| A2.11 | Psych stay in contact, if a pat is already registered for AS | Some pat are already registered with right-to-die organizations or are prepared for AS when psych discuss AS with them. Psych are also conversation partners on this topic in such cases. | The other was a phone call with a pat who is really severely limited in his quality of life [...] And he also said that he had joined a right-to-die organization because at some point, he wants to decide for himself when to end his suffering. (T09, 24) | 7 | 16 |
| **A3** | **Psych take on further tasks related to AS** | | | | |
| A3.1 | Psych support pat relatives during and/or after AS process | Psych support pat relatives, as AS can also be a significant burden for them. They support relatives of pat who want or have already realized AS. | We always feel responsible for the patients and their immediate, close social environment. [...] That means we take care of the year after. And I always have relatives who come afterward. There aren't many, but for some, maybe just the offer is helpful when we say, 'Feel free to reach out again after the funeral. Then we can see how you start your new life and what kind of help you can seek.' (T06, 56) | 5 | 8 |
| A3.2 | Psych organized AS-related professional events | Psych organize courses on AS at universities or offer training/workshops on AS or communication skills. | I also have some involvement with the topic in my role as a lecturer at the university, where we occasionally hold events that discuss the underlying issues | 4 | 4 |

| | | | | | |
|---|---|---|---|---|---|
| | | | surrounding these questions: whether one should be for or against it, and what role psychologists could play in this. (T06, 16) | | |
| A3.3 | Psych actively seek education and training on AS for themselves | If psych want to be well-informed and trained on AS, they must take the initiative to seek education, attend training, and engage in discussions. | Yes, it's not as publicly accessible as I would sometimes like it to be; rather, one must actively seek it out. [...] It's not something that's thrown at you everywhere; you're somewhat shielded from it unless you actively decide to engage with it, and then you can find information. (T04, 84) | 2 | 3 |
| **A4** | **Knowledge and perceived competence regarding AS vary among psych** | | | | |
| A4.1 | Psych feel inadequately informed and unprepared regarding AS | Psych are insufficiently informed regarding AS (e.g., terminology, legal framework, procedures with AS organizations, psychotherapeutic approaches, documentation of AS requests) and therefore inadequately prepared for AS tasks (e.g., counseling, assessing free decision-making). Reasons for the lack of knowledge include a lack of information, experience, coverage in training, AS-related training, and guidelines. | I believe I am still not well-prepared for almost all tasks. This includes having and being able to share information, such as what the current status is, what organizations are available, and all the practical details, as well as the therapeutic tasks. I think that in the way I approach it, it's fine, but I still feel very unsure therapeutically. I also wonder if there are specific therapeutic options or strategies? What should one definitely ask in order to help the person make a good decision for themselves? (T03, 76) | 9 | 40 |
| A4.2 | Psych feel insecure handling AS requests from certain vulnerable groups | Psychologists are uncertain about how to respond to AS requests from certain vulnerable groups. Examples include individuals with depression, intellectual disabilities, or physical impairments, children, or people without family support. | We've often had discussions within the team about the extent to which a wish for assisted suicide from someone with severe depression should be followed. What about people with intellectual disabilities? What about those with severe physical impairments who may not even be able to express themselves in full sentences? How can we ensure that these individuals are truly understood, so that assisted suicide can be supported in such cases? (T08, 44) | 3 | 4 |
| A4.3 | Psych feel well-prepared for aspects of AS | Some psych feel adequately prepared for certain tasks related to AS (e.g., knowledge about AS, psychotherapeutic interventions, support in decision-making, assessment of free decision-making). Contributing factors include professional experience, participation in seminars/trainings, or good collegial exchange. | Personally, I would feel quite well prepared now, but I also attend trainings offered by the German Society for Palliative Medicine and read current literature on the topic. [...] Without that, I wouldn't feel as confident. (T12, 77) | 8 | 12 |
| **A5** | **Legal and institutional frameworks influence the AS-related tasks of psych** | | | | |
| A5.1 | AS tasks are highly setting- and context-dependent | Requests AS vary depending on the setting (e.g., acute hospital, outpatient psychotherapy). This affects how often AS is addressed and whether discussions or counseling are possible. | But that is certainly also due to my specific work context. I think in a clinic or a counseling center – there are so many different formats – it might look different, | 12 | 40 |

| | | | | | |
|---|---|---|---|---|---|
| | | Psych may take on different roles depending on the context. For example, pat in acute hospitals often have stays too brief for support during the decision-making process. Institutional factors, such as religious affiliation and internal policies, also play a key role. | because the expectations placed on one another by the various institutions may differ. I may be more clear-cut in my role and know what I do and what I wouldn't do, also in terms of time constraints. But I believe that's a bit different in every work context. (T04, 88) | | |
| A5.2 | Guidelines and policies are unclear or lacking | Guidelines and regulations regarding AS are unclear for psych, both at the societal-legal level and within institutions. Key questions – such as the employer's stance on AS, what is permitted or expected, how processes and counseling should be structured, and what role psych should take – often remain unanswered. There is a lack of legal frameworks, institutional structures, clear directives, SOPs, QM documents, guidelines, and documentation standards. | "Right now, it's kind of like the Wild West regarding what is or isn't possible with AS. Due to the lack of legal regulations or the absence of a concrete draft law at the moment, I can well imagine that the number of requests will also increase. And here at [the clinic], we barely have any guidelines. There's no SOP, nothing in the quality management manual. We're really still completely starting from scratch." (T02, 38) | 9 | 23 |
| A5.3 | Institutional policies restrict AS work | Some hospitals prohibit any discussion of AS at the patient's bedside. This restricts Psych in their communication and actions, at times forcing them to act against their own convictions. Such restrictions are particularly common in religiously affiliated institutions. | What adds to it is that we are a Christian institution. So even talking about it at the patient's bedside is something that could lead to a formal warning (T08, 16) | 1 | 6 |
| A5.4 | The extent of collegial exchange varies | | | | |
| *A5.4.1* | *Collegial exchange on AS is limited* | Psych rarely or only partially discuss or document their experiences with AS among colleagues. | It's not something that gets widely shared, because we're not really sure what's legal and what isn't. For example, I wouldn't put that in writing to anyone. That's also why I don't hear much about it – it's just not something that circulates widely." (T01, 36) | 5 | 5 |
| *A5.4.2* | *Collegial exchange on AS occurs* | Psych have opportunities for collegial exchange on AS. For example, with open-minded colleagues or at conferences and professional events. | But I'm in weekly exchange with our palliative care department, which means I hear about it relatively quickly if something happens – if something is interesting or important. (T04, 80) | 4 | 5 |
| A5.5 | Suitable spaces are lacking | In hospital or acute care settings, undisturbed spaces for private conversations about AS with pat are often lacking | Structurally, this is unfortunately a major problem, because we have no dedicated space to offer private conversations on such sensitive topics. […] This is extremely unfortunate for a process that follows clearly defined stages and can be disrupted or interrupted by external disturbances. (T05, 76) | 1 | 1 |
| A5.6 | There is a lack of financial reimbursement for some AS-related tasks | Depending on the professional setting, some AS-related tasks cannot (easily) be reimbursed financially for psych. | If I'm only available in the inpatient setting and have already built a relationship there, I'm not allowed – due to billing regulations – to offer outpatient follow-up sessions. I see this as a serious risk for many patients | 2 | 2 |

| | | | | | |
|---|---|---|---|---|---|
| | | | who struggle to build trust quickly or to address sensitive issues. (T05, 48) | | |
| **RQ 2: What roles or tasks should psycho-oncologists undertake in requests for assisted suicide? (future directions)**<br>*[Note: Participants reported roles and tasks that they already undertake and future ones. Those corresponding to the current state are marked accordingly in the following.]* | | | | | |
| **B1** | **AS tasks must be based on the voluntariness of psych** | Psych involvement in AS-related tasks must remain voluntary and free from legal or employer-imposed mandates, respecting individual boundaries and possible conscientious objections of psych. | What I would find problematic is if it were a task that you couldn't refuse – if it weren't based on voluntariness. That would be an absolute no-go for me. The task and kind of support must be based on absolute voluntariness. (T01, 88) | 5 | 8 |
| **B2** | **Uncontroversial roles and tasks of psych regarding AS** | | | | |
| B2.1 | Psych should be open conversation partners *[already current state]* | Psych should serve as open conversation partners about e.g. suicidal thoughts, AS requests, and related topics, offering neutral support throughout this difficult decision-making process. What distinguishes psych from other professional groups is their stance: in conversations, they can adopt an open-ended, therapeutic, and respectful approach. | I believe the core and heart of it is what we, as psycho-oncologists, can provide, especially our attitude of not judging [...] that we clearly do not make decisions for someone else, but are happy to stand by their side when someone wants to make a difficult decision for themselves and seeks support in doing so. (T05, 92) | 8 | 17 |
| B2.2 | Psych should be "bridge builders" *[already current state]* | | | | |
| *B2.2.1* | *Psych should involve relatives* | Relatives should be included in the AS decision-making process, and the topic should be made discussable with them so they can understand the patient's reasoning and decision. Moreover, relatives should be considered as part of the patient's broader decision-making context. | Ideally, the family could be included, so that everyone is okay with it – or at least able to understand why it's so important to the person. That would really be ideal, if all emotionally involved individuals could be included and supported in the best possible way. Because suicide is never a one-person decision – well, in the end, unfortunately it is – but others inevitably get pulled in. And I find it very important, appropriate, and meaningful when others can also be part of that decision-making process. (T01, 60) | 6 | 10 |
| *B2.2.2* | *Psych should cooperate with other professionals* | Psych should not support pat with AS requests on their own but rather cooperate with other professional groups as part of a multidisciplinary team. These may include, in addition to psycho-oncologists, chaplains, physicians (e.g., psychiatrists, palliative care specialists, neurologists, and specialists for the respective disease group), nursing staff, psychotherapists, or palliative care psychologists. | It's definitely important that more than one professional group is involved. And this becomes especially important when the reasons or burdens leading to assisted suicide are primarily psychological rather than somatic. For example, as a psycho-oncologist, I would feel uncomfortable conducting the initial counseling for a patient with a neurodegenerative disease. (T02, 58) | 4 | 13 |

| | | | | | |
|---|---|---|---|---|---|
| B2.3 | Psych should explore AS wishes *[already current state]* | Psych should explore AS requests to support the patient's decision-making process. In-depth psychological exploration can facilitate well-considered choices by addressing motivations, concerns, social context, resources, and relevant life experiences. | First, we need to start by exploring these questions, these wishes. (T08,52) | 7 | 17 |
| B2.4 | Psych should provide neutral information about AS *[already current state]* | Psych should provide nonjudgmental information about AS and educate pat accordingly. To do so, Psych themselves must be sufficiently informed and prepared. | Well, ideally, it's helpful to provide information in advance when someone comes with such thoughts – about what assisted suicide looks like. Offering psychoeducational guidance, that would be the kind of preparatory work. (T09, 48) | 5 | 7 |
| B2.5 | Psych should discuss alternatives to AS *[already current state]* | Psych should discuss alternatives to AS with pat. For example, by exploring other treatment options (e.g., palliative care) or ways to improve their quality of life. | On the one hand, what need or distress is behind the thoughts of assisted suicide – and is it possible to work on that preventively, so the person no longer sees the necessity. (T07, 105) | 2 | 3 |
| B2.6 | Psych should refer pat to additional support structures *[already current state]* | Psych should be informed about appropriate points of referral (e.g., right-to-die organizations, psychotherapists, oncologists) and be able to direct pat or relatives accordingly. | That is definitely part of our role as psycho-oncologists: to be a point of contact, to provide information, and then to be able to refer further. (T11, 64) | 7 | 9 |
| B2.7 | Psych should offer psychotherapeutic interventions *[already current state]* | Psych should offer psychotherapeutic interventions when needed. | And possibly also to incorporate psychotherapeutic interventions. (T08, 72) | 3 | 4 |
| B2.8 | Psych could be involved early in someone's AS considerations *[additional future role/task]* | Psych should be approachable on the topic of AS early on, not only in late stages of illness or palliative situations. | But I think that for many people, the engagement with the possibility of assisted suicide begins much earlier in the course of the illness. And I find that currently, there are few people available to talk to about it. […] Also, given our training, we should be available to discuss such topics – even if they arise early in the course of the illness. (T12, 53) | 1 | 1 |
| B2.8 | Psych could address spiritual and religious concerns *[additional future role/task]* | Spiritual and/or religious concerns should be explored and integrated by psych in conversations about AS. | And that questions of faith are included. I've heard things here from deeply religious Christians like, 'I just can't go on – what happens if I take my own life, would God be okay with that?' And discussing such questions is part of the psychological domain. (T08, 48) | 2 | 2 |
| B2.9 | Psych could offer supervision on AS to other health care professionals *[additional future role/task]* | Psych consider it important that health care professionals involved in AS have access to supervision or collegial exchange to process their emotional responses. Psych could offer AS- | Whether it's psycho-oncologists or not, if someone offers supervision in this area, I think they need to have a relevant background. But whether that comes with the | 11 | 15 |

| | | | | | |
|---|---|---|---|---|---|
| | | related supervision to other psych, psychologists, or other specialties. | WPO [German professional society for psycho-oncology] certificate or not, I believe each team has to decide for themselves. Still, there has to be a qualified offer. (T04, 76) | | |
| **B3** | **Controversial roles and tasks of psych regarding AS** | | | | |
| B3.1 | Assessment of free decision-making<br>*[already current state]* | | | | |
| *B3.1.1* | *Psych should assess free decision-making* | Assessing free decision-making could be a task for psych. For example, by evaluating whether a mental disorder is influencing the AS request. The assessment and the support during the decision-making process should be carried out by different psych. | What speaks in favor of involving our professional group is that we are the experts in solid psychological diagnostics. Just yesterday in supervision, a supervisee told me that on the oncology ward where she works, physicians immediately diagnose a severe depression when someone talks about assisted suicide. That's a clear example where our expertise lies – in assessing the psychological state and properly evaluating free decision-making. I see a strong reason why we, as a psychological profession, should take an active role here. (T12, 49) | 9 | 19 |
| *B3.1.2* | *Psych should not assess free decision-making* | Some psych reject the assessment of free decision-making as part of their role, viewing psychiatrists or right-to-die organizations as more appropriate for this task. They do not see themselves in an evaluative or expert-review role. | I myself am very cautious, because I really don't want to take on that kind of evaluative or expert role. And I assume that if someone joins a right-to-die organization, there are assessment procedures in place there. (T09, 28) | 5 | 8 |
| B3.2 | Psych should support pat during AS planning<br>*[already current state]* | Psych may occasionally be involved in the planning of AS – for example, in clarifying who could or should be present or provide support. However, the planning of AS is primarily regarded as a medical responsibility. | I would assume that questions like which medication to use need to be explained by a physician. But of course, when it comes to who in someone's social environment might be suitable, whom they could ask, and how to go about it – that's where, in my context, I would always offer to have conversations with one or two of the people involved, in pairs or small groups, whatever fits. (T04, 56) | 3 | 5 |
| B3.3 | Support of pat relatives during and/or after AS process<br>*[already current state]* | | | | |
| *B3.3.1* | *Psych should support pat relatives* | Relatives should have access to support from Psych, including pre- and post-AS care. | [...], but above all, that the relatives, the companions, or the assisting persons know: once it's formally over – once the person has died – there's still a safety net. If | 10 | 13 |

| | | | | | |
|---|---|---|---|---|---|
| | | | something suddenly arises that burdens me, I have relatively quick access to support. (T04, 72) | | |
| *B3.3.2* | *Psych should not be responsible for pat relatives* | Psych should not be responsible for supporting pat, including pre- and post-AS care. | Personally, I'd rather not have my calendar packed with grieving relatives after an assisted suicide (T02, 54) | 1 | 1 |
| B3.4 | Mandatory psychological counseling regarding AS<br>*[additional future role/task]* | | | | |
| *B3.4.1* | *Psych should provide mandatory counseling on AS* | Psych should conduct at least one mandatory counseling session in response to AS requests, as they are particularly well suited for this task. | As things stand, I would hope for something like a mandatory counseling requirement. And maybe also not just one session, but two – spaced a few weeks apart – to help prevent impulsive decisions made in a moment of emotional distress. (T06, 48) | 3 | 4 |
| *B3.4.2* | *Psychologists reject mandatory counseling on AS* | Psych reject mandatory counseling for pat regarding AS, as it creates poor conditions for an open-ended conversation. | I think it's a different matter if, in a future legally regulated process, this becomes our defined task. Then it immediately has the downside of being a mandatory appointment that patients have to attend in order to get the certificate. The downside is – I always find mandatory appointments problematic. They're always a poor foundation for an open-ended conversation. (T12, 57) | 1 | 3 |
| B3.5 | Involvement during realization of AS<br>*[additional future role/task]* | | | | |
| *B3.5.1* | *Psych could support pat throughout the entire AS process, including realization* | In individual cases, psych could provide psychological support throughout the entire AS process, including during the act itself, if supported by the therapeutic relationship and desired by both pat and psych. However, concrete implementation strategies would first need to be developed. | For these individuals, I would definitely wish for at least the option of psychotherapeutic or psycho-oncological support. At that point – once it's been determined that the person can go through with it – maybe a psych or someone in that direction could accompany them. The question isn't just how it's done, but also how dying is shaped and supported, and how someone stays in contact during that. How can we make it a dignified process? (T11, 52) | 2 | 3 |
| *B3.5.2* | *Psych should not be involved during realization of AS* | Psych should not be involved in the actual act of AS, as this is generally regarded as a medical responsibility. | As for accompanying someone during assisted suicide – I don't think everyone would want to do that. And for that reason, I don't think it should be a task assigned to psych. (T01, 56) | 7 | 8 |
| **B4** | **Necessary competencies and involvement of other professions** | | | | |

| | | | | | |
|---|---|---|---|---|---|
| B4.1 | AS conversations partners should have appropriate and sufficient competences regarding AS | AS-related conversations should be conducted by specially trained and competent professionals. It is important that they have psychotherapeutic training, are well-informed about AS, and have sufficient professional experience. These individuals may, but do not necessarily have to be psych. | You need to undergo further training. I think this topic deserves to have people, who are specifically trained for it. (T06, 52) | 8 | 20 |
| B4.2 | Other professionals are also suitable for AS talks | Psych are not the only professionals suited for certain AS-related tasks; these can also be carried out by other professional groups. This includes, for example: aftercare for relatives (e.g., grief counselors or structured group programs); AS-related conversations (e.g., physicians, night nurses, social environment, disease-specific specialists, psychotherapists); and presence during the act of AS (e.g., hospice services, end-of-life companions). | Sometimes it's physicians whom patients have a good connection with, sometimes it's conversations with the night nurse if there's a strong rapport. And of course, some are so well supported by their own social environment that they can talk it through with them. There's no obligation to discuss it with a psycho-oncologist. But if there's no one else, and the psycho-oncologist is willing, then yes. I think it's good if our professionals are open to such conversations. (T06, 44) | 7 | 9 |
| B4.3 | Palliative care psychologists may be especially qualified for AS work | Psych identify palliative care psychologists as valuable points of contact for AS-related concerns. They are more specifically trained than psych and could, for example, take on counseling and/or support of relatives. | And as I said, when it comes to aftercare, yes, that could be our task. But as mentioned earlier, it might be something where palliative care psychologists are a better fit. Because they're not only familiar with dying in oncology, but also with cases where people have died from other causes or chosen assisted suicide due to other illnesses. (T02, 78) | 3 | 6 |
| **RQ 3: What do psycho-oncologists need to feel well-equipped to perform these roles and tasks? (needs)** | | | | | |
| **C1** | **Psych need training on and preparation for AS** | | | | |
| C1.1 | Psych need specific training on AS | To be well prepared for working with AS requests, psych express a need for education and training opportunities on the topic. These could be organized as standalone AS-focused courses or be integrated into existing psycho-oncology programs. Desired content includes legal frameworks, therapeutic approaches to assessing free decision-making, and procedures related to the implementation of AS. | And I would wish [...] that in the future, for the people conducting this counseling, we organize training sessions with materials to enable them to carry out this counseling responsibly. For example, how can one assess free decision-making? How can one determine whether there is social pressure involved in the decision-making process? (T06, 48) | 11 | 29 |
| C1.2 | Psych need practical workshops on AS communication skills | Psych need practice-oriented workshops in communication to be prepared for AS-related conversations in clinical practice. Experienced Psych could design and lead these seminars. | What I find really problematic is that, due to time constraints, practical training in communication skills is offered at very few universities. […] And in further training, there's no practical practice either. Various therapeutic tools are discussed. But how does it actually work in practice? My colleagues are figuring that out on their own. (...) It would be important to have training seminars, especially on assisted suicide. What do I need | 1 | 3 |

| | | | | | |
|---|---|---|---|---|---|
| | | | to pay attention to? What should such conversations include? (T08, 88) | | |
| C1.3 | Psych need clarity about their personal attitudes toward AS | AS is a controversial topic that touches on deep personal convictions. Psych should engage with it early and reflect on their own stance (e.g., through self-exploration) to remain open-ended in therapy. | And that there should be more elements of self-exploration. I think you first need to form your own opinion or attitude – how you personally feel about it – and make sure that this attitude doesn't influence the therapeutic contact. Just because I personally decide [...] that I don't think it's a good thing, I would still have to counsel and inform patients properly. (T03, 88) | 5 | 6 |
| **C2** | **Psych need support and self-care** | | | | |
| C2.1 | Psych need supervision and collegial exchange | Psych need regular, field-specific supervision, quality circles, or collegial exchange to manage the high emotional strain and personal impact associated with the topic of AS. | To do this kind of work, I would need a strong team, good supervision, and collegial exchange. I would also need someone who has already done it and could support me with advice – someone who can speak from personal experience. That would be very helpful. (T01, 80) | 8 | 16 |
| C2.2 | Psych need self-care | Psych must be able to engage in recognized self-care. They need sufficient space to attend to their own needs in order to maintain psychological stability. | And we psychologists need something like: 'I just had a terrible, heavy counseling session. Give me an hour of peace, even if lunch break is already over.' That doesn't work like in rehab, where secretaries completely overbook psychologists – half an hour, a rushed lunch. No. This emotional burden has to be manageable. (T08, 72) | 3 | 4 |
| **C3** | **Psych need evidence-based guidelines and tools** | Psych need evidence-based guidelines and tools to navigate AS-related practice. These should include procedures for assessing free decision-making, counseling approaches, and documentation standards. In addition, psych require informational materials that can be shared with pat. | I would also like to have a guideline, a brochure from WPO [German professional society for psycho-oncology] or whatever, that outlines: What alternatives to assisted suicide can I present in psychoeducational conversations? What [assisted suicide] associations exist? How do they typically work? What are the legal foundations? A brochure like that would provide good orientation and could be something to share with patients. And a guideline specifying which key areas should definitely be addressed in conversations (T09, 60) | 9 | 20 |
| **C4** | **Psych need institutional and healthcare system changes** | | | | |
| C4.1 | Institutional policies should not restrict open AS conversations | Institutional policies should not prevent psych from having open and practically helpful conversations about AS with pat. | Quite often, I find myself wishing it were clearer that we are allowed to include these topics in our conversations. (T05, 36) | 2 | 2 |

| | | | | | |
|---|---|---|---|---|---|
| C4.2 | Psych need a well-organized team with good communication | Working with AS requires a strong team and effective communication among all treating professionals. | First, good team communication. And in acute care settings, it's still a big issue that many physicians have no real understanding of what psychologists actually do. That's still the case." (T08, 76) | 5 | 5 |
| C4.3 | Psych need confidential spaces for AS conversations | Psych need confidential spaces where they can talk privately with pat about AS. | Second, I need a space where I can do this in peace, without interruptions – where no one keeps bursting in. Where it's not like, 'I need to draw blood now,' or 'he has to go to the ECG,' or whatever else. That would be really important. A space like a patient room. I press a red button, and no one comes in anymore. (T08, 68) | 3 | 6 |
| C4.4 | Psych need adequate time for pat interactions | Conversations about AS or end-of-life wishes are time-intensive and must be institutionally supported and enabled. | Of course, the time I have with such a person is much too short for that. I would think that enough time and in-depth conversation spaces are needed to accompany such matters more thoroughly with information. (T11, 40) | 4 | 7 |
| C4.5 | AS-related tasks must be eligible for financial reimbursement | AS-related tasks must be financially covered to enable psych to take them on. This includes general financial barriers, such as billing for post-discharge contacts, as well as the specific funding of AS-related activities. It is also essential that psych remain financially independent from the execution of AS to maintain professional neutrality. | Ideally, it would just be a completely standard health insurance service. A counseling session on this topic could be billed just like a palliative care consultation. It's actually equivalent, one would think, but I believe there are still a few hurdles to overcome. (T05, 40) | 6 | 12 |
| C4.6 | Psych need clear referral pathways and contact persons responsible for AS | Clearly designated AS contact persons or services are needed, to whom psych can refer and who serve as points of contact for pat. They should offer structured spaces for discussion (e.g., AS consultation hours, specialized AS teams). | At the same time, maybe within the whole hospital, clinic, or wherever one works, there should be some reflection on who can take responsibility for this. Or the development of a guideline indicating who to turn to, even within the clinic. The key is having clearly identified individuals or those willing to engage with the topic, so that everyone knows, and referrals can be made to the right people. (T10, 92) | 6 | 11 |
| C4.7 | There should be AS-related information sessions for pat | There should be informational events for pat on the topic of AS, followed by the option to receive further counseling. | What I would really appreciate is a combined offer of informational events. It sounds odd, but something like a 'Day of Assisted Suicide', where various aspects are presented from different professional perspectives through lectures or perhaps a panel discussion. So people can hear different viewpoints, without needing to have a formed opinion themselves and be supported in their own decision-making process. And then have the option to register for one-on-one sessions, held in a | 1 | 3 |

| | | | | | |
|---|---|---|---|---|---|
| | | | classic setting with an appropriate room and protected conditions. (T05, 40) | | |
| C4.8 | Psychologists need clear legal regulations | Psych wish for clear legal regulation of AS that protects healthcare professionals – now and in the future – from legal uncertainty. Legal clarity would also help pat understand what is possible and how. The regulation should strike an appropriate balance between autonomy/self-determination and the protection of life. Access criteria for AS must be clearly defined. | And as for the lawmakers, things still aren't clearly defined; it's all a bit vague. I believe it would be important for there to be really concrete guidelines and recommendations, or however you want to put it. That it is all very specific, so that one doesn't have to be afraid of facing difficulties when doing this or helping with it [assisted suicide]. There was that case in Berlin with the psychiatrist who was convicted. That's exactly why I think it needs to be clarified further – because if you hear things like that, you're more likely to keep your distance from the topic if there's a risk it could come back to harm you (T10, 96) | 6 | 8 |
| C4.9 | Psych need societal acceptance of AS | Psych wish for greater societal acceptance, understanding, recognition, and goodwill toward AS and their work in this area. AS should become a topic that can be openly discussed and destigmatized in society. | I think it's important to give this topic a great deal of space and attention, because it's ultimately about life and death. And I also believe it's essential to make it discussable and to take it out of this forbidden zone, because it's a reality that exists. I hope that by destigmatizing the topic, we might reduce suicides – accidental suicides, or I'm not sure what to call them – but suicides as they currently still mostly happen. (T01, 92) | 5 | 7 |
